# A View-Agnostic Deep Learning Framework for Comprehensive Analysis of 2D-Echocardiography

**DOI:** 10.1101/2025.07.10.25331304

**Authors:** D M Anisuzzaman, Jeffrey G. Malins, John I. Jackson, Eunjung Lee, Jwan A. Naser, Behrouz Rostami, Jared G. Bird, Dan Spiegelstein, Talia Amar, Christie C. Ngo, Jae K. Oh, Patricia A. Pellikka, Jeremy J. Thaden, Francisco Lopez-Jimenez, Timothy J. Poterucha, Paul A. Friedman, Sorin V. Pislaru, Garvan C. Kane, Zachi I. Attia

## Abstract

Echocardiography traditionally requires experienced operators to select and interpret clips from specific viewing angles. Clinical decision-making is therefore limited for handheld cardiac ultrasound (HCU), which is often collected by novice users. In this study, we developed a view-agnostic deep learning framework to estimate left ventricular ejection fraction (LVEF), patient age, and patient sex from any of several views containing the left ventricle. Model performance was: (1) consistently strong across retrospective transthoracic echocardiography (TTE) datasets; (2) comparable between prospective HCU versus TTE (625 patients; LVEF *r*^2^ 0.80 vs. 0.86, LVEF [> or ≤40%] AUC 0.981 vs. 0.993, age *r^2^* 0.85 vs. 0.87, sex classification AUC 0.985 vs. 0.996); (3) comparable between prospective HCU data collected by experts versus novice users (100 patients; LVEF *r^2^* 0.78 vs. 0.66, LVEF AUC 0.982 vs. 0.966). This approach may broaden the clinical utility of echocardiography by lessening the need for user expertise in image acquisition.

## Introduction

Echocardiography stands as a fundamental diagnostic tool in cardiology, traditionally reliant on the meticulous selection of specific viewing angles and the interpretative expertise of human operators. This conventional approach, while effective, harbours inherent limitations due to its dependency on the skill and experience of the echocardiographer. For example, certain views are particularly challenging for inexperienced operators to acquire and select ^1^. As a result, the conventional approach potentially leads to variability in assessments and constraints in the scope of analysis ^2^. Moreover, due to a shortage of trained sonographers globally ^3^, significant delays in obtaining a transthoracic echocardiogram (TTE) exist at many medical centres. In response to these challenges, this study presents a view-agnostic deep learning framework for echocardiographic analysis that reduces reliance on specific views.

A view-agnostic learning framework may be especially useful for handheld cardiac ultrasound (HCU) images. HCU, which when deployed in point-of-care settings such as patient bedside is termed point-of-care ultrasound or POCUS, typically focuses on clinically relevant questions regarding the presence or absence of specific cardiovascular abnormalities and is often acquired by individuals with modest training. This variability in operator experience can translate to an even greater possibility for variability in image quality and study interpretation compared to a standard TTE, and consequently a greater need for a robust framework that is less susceptible to this potential variability.

The objective of this study is to introduce and validate an imaging approach that eliminates the traditional requirements of view-specific echocardiography, enabling acquisition of data by users with minimal training. We hypothesized that by leveraging the capabilities of advanced convolutional neural networks trained using multiple TTE views, we could analyse echocardiographic data (either TTE or HCU) from any 2D view designated as valid (that is, containing the left ventricle). To test this hypothesis, we employed a range of retrospective and prospective echocardiographic datasets to develop, train, and validate a set of view-agnostic models. The first model demonstrates the power of this approach in estimating left ventricular ejection fraction (LVEF) – a basic measurement in echocardiography – whereas the second and third models respectively estimate a patient’s age and sex from echocardiographic images, thereby further demonstrating the power of this approach in extracting physiologic information.

## Methods

### Data acquisition and selection

All study procedures were approved by the Mayo Clinic Institutional Review Board, and we have complied with all relevant ethical regulations. Patient cohorts only included individuals who had provided prior authorization for the use of their data in research.

The model training cohort consisted entirely of TTE patients from Mayo Clinic Rochester and surrounding Mayo Clinic Health System sites in Minnesota and Wisconsin. In this dataset, there was one exam per patient from 19,627 patients total, with all echocardiogram study dates between January 1, 2007 and September 30, 2022. Each patient was randomly assigned to either the training, validation, or testing datasets to avoid cross-contamination of the datasets. The validation dataset was used to make decisions regarding model architecture as well as hyperparameter tuning.

Clinically indicated echocardiography was recorded in clips of three cardiac cycles and performed using one of the following machines: GE Healthcare E95 or S70, Philips Epiq, or Siemens Acuson Sequoia. The reference measured LVEF was based on the clinical measure using the following hierarchy ^4^: 3D volumetric measure, followed by a 2D biplane measurement (apical views), 2D modified Quinones, M-mode modified Quinones, and lastly, a 2D visual estimate by a level III trained expert echocardiographer.

In addition to the randomly selected set of patients from Minnesota and Wisconsin that we used for internal testing, we also evaluated models using several additional testing datasets, as described below. More details regarding inclusion and exclusion criteria can be found in the Supplementary Material.

1. Randomly selected patients with TTE exams collected at the Mayo Clinic site in Scottsdale, Arizona (1,695 patients) between January 1, 2022 and February 29, 2024
2. Randomly selected patients with TTE exams collected at the Mayo Clinic site in Jacksonville, Florida (1,862 patients) between January 1, 2022 and February 29, 2024
3. The EchoNet Dynamic dataset, which is a publicly available dataset of TTE clips from 10,030 patients collected at Stanford University Hospital (data from 10,015 patients used for analysis; one A4C clip per study). Use of this dataset complies with the Stanford University School of Medicine Research Use Agreement and Terms of Use.
4. A prospective cohort of 625 patients who had TTE and HCU data collected during the same session. These patients visited Mayo Clinic Rochester for a clinically indicated TTE exam between November 1, 2022 and September 30, 2023. Immediately following their TTE exam (i.e., within the same session), if patients provided verbal consent for additional clips to be collected for research purposes, the following five HCU 2D video clips were collected using a Philips Lumify device: PLAX, parasternal short axis, A4C, A3C, A2C.
5. An HCU dataset of 100 patients collected across three external sites (35 patients from Aurora St. Luke’s Medical Center in Milwaukee, WI; 32 patients from the University of Chicago Medical Center; 33 patients from Sheba Medical Center in Israel). The study protocol was approved by each respective institution’s IRB committee, and all patients were consented according to the study protocol. From each patient, two operators each collected up to 10 views of the heart using a Philips Lumify device. One of the operators was an experienced sonographer, whereas the other operator was one of 9 novice users who used real-time guidance AI software (UltraSight Ltd.) to optimize the acquired views ^1^. From the full exam, if more than one of the A2C and A4C views were obtained, the highest quality clip was selected for each view, as determined by the same real-time guidance AI software used for acquisition (UltraSight Ltd.).

### Data preprocessing

DICOM (Digital Imaging and Communications in Medicine) data were preprocessed using a framework previously described ^5^, which included steps such as identification of the echocardiographic imaging sector, removal of portions of the electrocardiogram trace overlapping this sector (using *opencv* version 4.5.5 in Python ^6^), and view classification of video clips. Clips were included for analysis only if they were B-mode, had at least 48 frames, and belonged to one of four ‘valid’ view categories containing the left ventricle: parasternal long axis (PLAX), apical 2-chamber (A2C), apical 3-chamber (A3C), and apical 4-chamber (A4C). Figure 1 provides an overview of the data processing workflow. More details regarding the preprocessing workflow can be found in the Supplementary Material.

**Figure 1:**
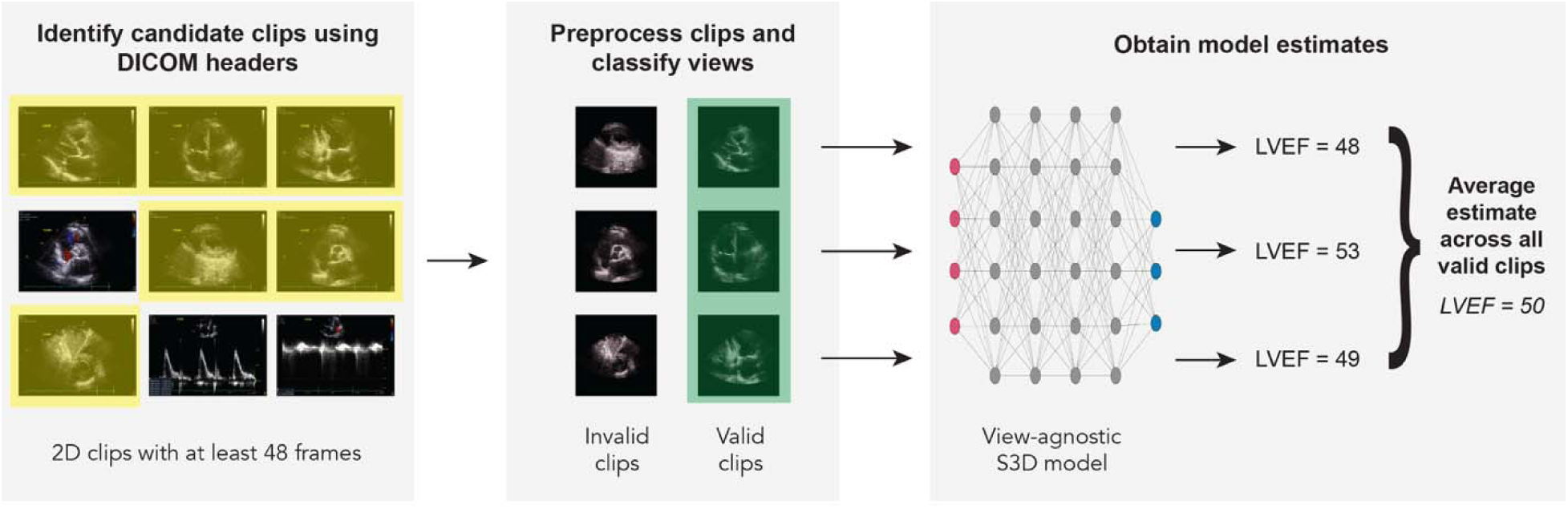
An overview of the view-agnostic deep learning framework employed in the current study. Note that the LVEF model estimates are hypothetical and do not constitute actual data points from the study. The icon used to denote the deep learning model was taken from BioRender.com.

### Model training

We used the S3D model architecture developed by Xie et al. ^7^, but adapted it to take in single channel (grayscale images) input. We first trained a model for LVEF estimation from scratch (i.e., without using the pre-trained weights for the S3D model) and saved the best model based on validation loss. This saved model’s weights were then used for transfer learning to train age estimation and sex classification models.

We selected 24 frames to constitute a single input video by sampling every other frame from a fixed-length segment. To increase model generalizability, we applied five augmentations to the training dataset: random rotation between-10 and +10 degrees, Gaussian blurring, central and random cropping, and horizontal flipping ^8^. With temporal sliding windows and augmentations, the models were trained on 473,803 clips from 19,627 patients. More details regarding model training and selection can be found in the Supplementary Material.

### Evaluating model performance

To evaluate performance of the LVEF estimation model, in addition to assessing continuous output from the regression model, we also performed a binary classification between significantly reduced LVEF (clinically-calculated LVEF ≤ 40%) versus normal or mildly reduced LVEF (clinically-calculated LVEF > 40%). To do this, we first obtained the threshold LVEF value that balanced sensitivity and specificity for the validation dataset. We then applied this threshold (45.72%) to the testing datasets. For age, we performed a continuous regression, whereas for sex, we performed a binary classification between “male” and “female”.

For the retrospective TTE datasets, we averaged model estimates across all valid clips containing the left ventricle. For the prospective dataset of simultaneously collected TTE and HCU clips, we instead selected one clip per view for the TTE dataset (i.e., the clip with the highest view classifier inference score for each view ^9^) to enable a direct comparison with the HCU dataset, for which only clip per view was collected.

Details regarding the model evaluation metrics that we used can be found in the Supplementary Material.

## Results

### Sample characteristics

Patient demographics and clinical characteristics are presented in Table 1. As shown in Table 1, (1) mean LVEF was lower in the model development datasets compared to the model evaluation datasets because the model development datasets were specifically enriched for the middle-to-lower end of the LVEF distribution, and (2) demographic characteristics and comorbidities differed according to geographic location, and were also influenced by patient selection criteria for the various cohorts.

**Table 1.** Demographic and clinical characteristics of the patient cohorts for model development and model evaluation.

|  | Model Development<br>(Minnesota-Wisconsin) |  |  | Model Evaluation |  |  |  |
| --- | --- | --- | --- | --- | --- | --- | --- |
|  | Training | Validation | Testing | Arizona | Florida | TTE &<br>HCU<br>Collected<br>from Same<br>Patients | HCU<br>Collected<br>by Experts<br>and Novice<br>Users |
| Number of studies | 19,627 | 862 | 1,890 | 1,695 | 1,862 | 625 | 100 |
| Patient Demographics |  |  |  |  |  |  |  |
| Age on study date (years) |  |  |  |  |  |  |  |
| mean ± standard deviation | 64 ± 16 | 65 ± 16 | 66 ± 17 | 68 ± 15 | 66 ± 15 | 64 ± 16 | 60 ± 17 |
| range | 18-106 | 18-97 | 18-104 | 19-103 | 18-100 | 18-96 | 22-90 |
| Sex (%) |  |  |  |  |  |  |  |
| female | 45.21 | 41.42 | 43.28 | 40.94 | 45.38 | 39.52 | 38.00 |
| male | 54.78 | 58.58 | 56.72 | 59.06 | 54.62 | 60.48 | 62.00 |
| Race and ethnicity (%) |  |  |  |  |  |  |  |
| NH American Indian / Alaska Native | 0.55 | 0.23 | 0.37 | 1.77 | 0.16 | 0.48 | – |
| NH Asian | 1.35 | 1.86 | 0.79 | 3.78 | 2.26 | 2.08 | 2.00<sup>†</sup> |
| NH Black / African American | 2.06 | 1.51 | 2.12 | 2.95 | 9.77 | 2.40 | 22.00<sup>†</sup> |
| Hispanic / Latino | 2.16 | 2.44 | 2.38 | 9.26 | 5.69 | 1.44 | 6.00 |
| NH Native Hawaiian / Other Pacific<br>Islander | 0.06 | 0.12 | 0.16 | 0.35 | 0.21 | – | – |
| NH White | 89.23 | 90.37 | 90.63 | 79.41 | 79.16 | 91.20 | 76.00<sup>†</sup> |
| Other or unknown | 4.60 | 3.48 | 3.54 | 2.48 | 2.74 | 2.40 | – |
| Comorbidities (% positive) |  |  |  |  |  |  |  |
| Hypertension | 60.03 | 64.85 | 68.57 | 68.32 | 67.40 | 59.52 | 57.00 |
| Atrial fibrillation | 31.05 | 32.25 | 34.60 | 35.46 | 31.42 | 35.20 | 10.00 |
| Hyperlipidaemia | 55.43 | 57.19 | 62.22 | 62.77 | 61.17 | 58.72 | 48.00 |
| Congestive heart failure | 41.38 | 41.18 | 45.71 | 38.17 | 36.31 | 45.76 | 22.00 |
| Myocardial infarction | 18.31 | 18.56 | 23.60 | 20.00 | 18.26 | 12.00 | 20.00 |
| Valvular disease | 49.45 | 56.15 | 51.01 | 73.92 | 63.27 | 66.88 | NA |
| Peripheral vascular disorders | 39.05 | 44.20 | 44.23 | 44.42 | 40.66 | 50.56 | NA |
| Chronic pulmonary disease | 35.39 | 28.77 | 34.76 | 25.49 | 27.07 | 25.92 | NA |
| Diabetes mellitus | 25.26 | 23.67 | 28.68 | 23.95 | 27.77 | 19.04 | 23.00 |
| Renal failure | 27.22 | 24.59 | 32.12 | 31.74 | 31.20 | 27.36 | NA |
| Liver disease | 18.60 | 15.20 | 17.46 | 19.29 | 24.17 | 16.96 | NA |

| Echocardiography |  |  |  |  |  |  |  |
| --- | --- | --- | --- | --- | --- | --- | --- |
| Left ventricular ejection fraction* |  |  |  |  |  |  |  |
| mean ± standard deviation | 55 ± 12 | 54 ± 12 | 54 ± 13 | 59 ± 10 | 60 ± 10 | 57 ± 11 | 56 ± 12 |
| range | 6-87 | 15-74 | 10-79 | 12-79 | 6-82 | 11-75 | 13-78 |
| Equipment manufacturer (%) |  |  |  |  |  |  |  |
| GE Healthcare | 85.99 | 94.78 | 85.03 | 99.41 | 97.21 | TTE: 100 | – |
| Philips Medical Systems | 12.19 | 5.22 | 14.97 | 0.59 | 2.79 | HCU: 100 | 100 |
| Siemens Acuson | 1.82 | – | – | – | – | – | – |

*Note.* Sex as well as race and ethnicity were self-reported and obtained from each patient’s medical record. For all samples except for the external HCU dataset collected by experts and novice users (which had site-specific methods for defining comorbidities), comorbidities were ascertained using the *comorbidity* package in R <sup>37</sup> based on ICD-9 or ICD-10 codes noted in the patient’s medical record up to and including the echocardiographic study date. NH = non-Hispanic; NA = not assessed in this sample; A4C = apical 4-chamber; †Percentage includes patients who identify as Hispanic and those who identify as non-Hispanic, as race and ethnicity data were separately reported for the sample in aggregate
\*There was one patient in the training cohort whose LVEF was reported as 112, which we considered to be an error in the report. The mean and standard deviation values for LVEF in the training cohort are reported with this value taken out.

### LVEF estimation for TTE

We evaluated the LVEF regression model on TTE datasets from Minnesota-Wisconsin, Arizona, and Florida, as well as the publicly available EchoNet dataset (Figure 2). For Minnesota-Wisconsin, Arizona, and Florida we used all available valid-class clips, and so the result for each patient represents the average of the model outputs for each clip. In contrast, the EchoNet dataset contains only apical 4-chamber (A4C) clips, so there was only one single estimate from one clip per patient. Performance was strong in all cases, which was further confirmed by the Bland-Altman plots shown in Figure 3. As shown in Figure 3, for all four cohorts, the difference between model-estimated and clinically-calculated LVEF was less than 10% in at least 90% of cases.

**Figure 2:**
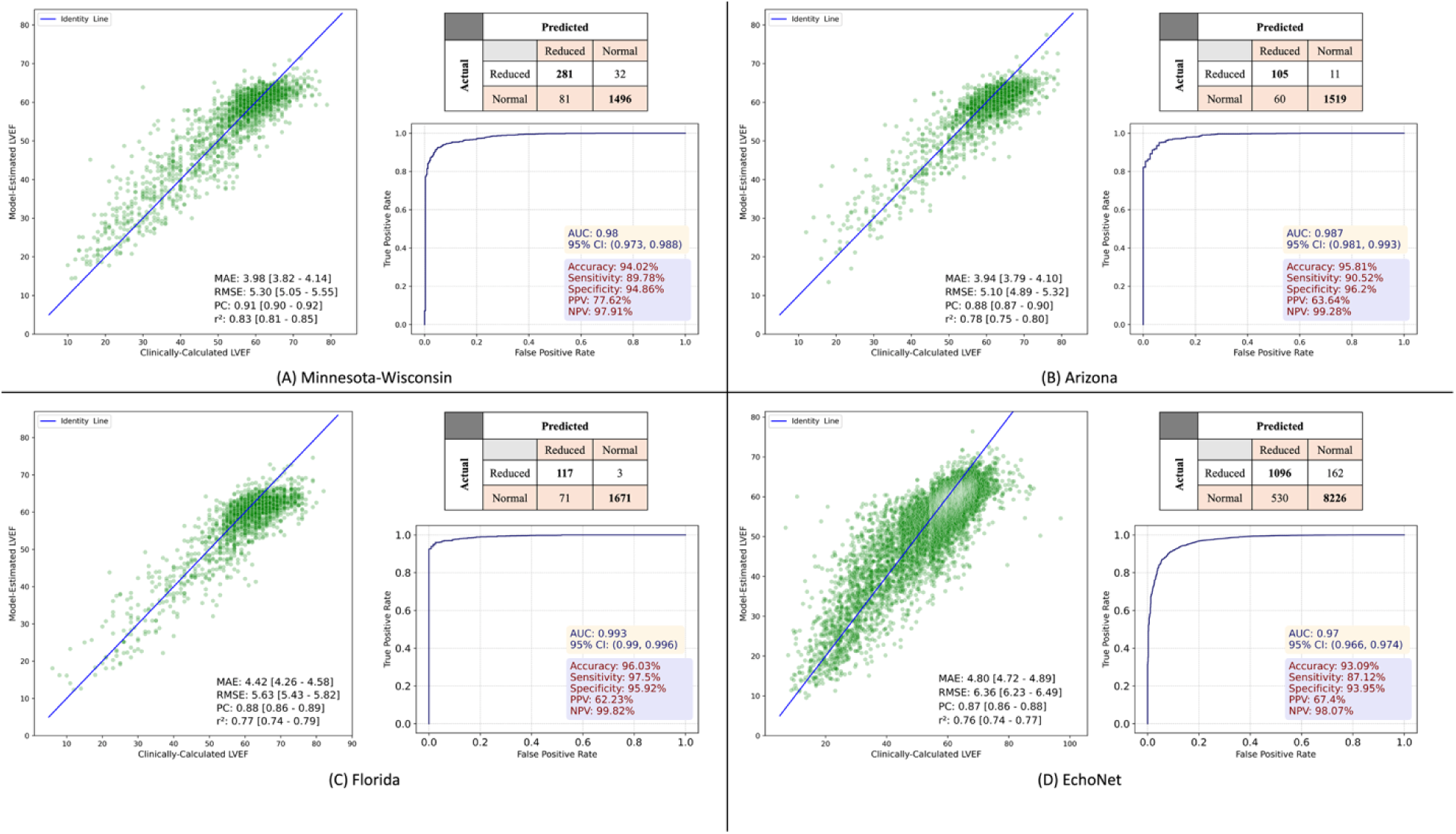
LVEF estimation and classification for the (A) Minnesota-Wisconsin, (B) Arizona, (C) Florida, and (D) EchoNet datasets. In each subplot, the left plot shows the correlation for LVEF estimation (i.e., comparison of model-estimated LVEF to clinically-calculated LVEF), whereas the right plot shows the ROC curve and confusion matrix for reduced versus normal LVEF classification.

**Figure 3:**
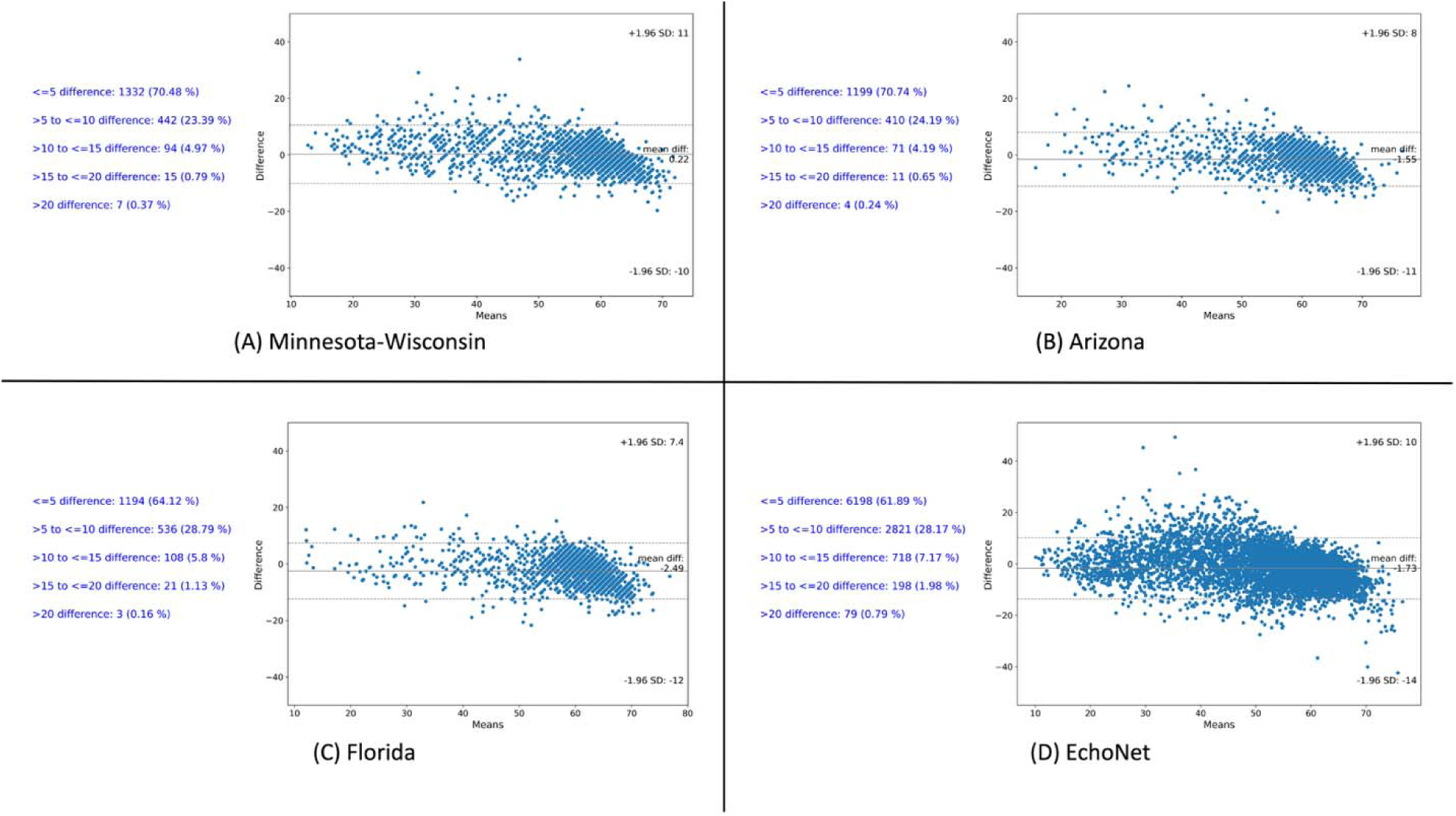
Bland-Altman plots showing the discrepancy between clinically-calculated and model-estimated LVEF for the (A) Minnesota-Wisconsin, (B) Arizona, (C) Florida, and (D) EchoNet datasets. The text in blue shows the number of cases with errors within certain bands (e.g., less than 5%, between 5% and 10%, and so on).

The decision to use all available valid-class clips was based on a preliminary analysis evaluating the LVEF regression model using different numbers of randomly selected clips and all available valid-class clips for the Minnesota-Wisconsin dataset (Figure 4). Experimental setups consisted of pulling between one and five random clips from all available valid-class videos for each patient and then averaging estimates across the clips that were pulled. This procedure of pulling random clips and averaging estimates was repeated 15 times, giving rise to the distributions shown in the leftmost five boxplots in each subplot. As shown in Figure 4, as the number of valid-class clips increased, the median RMSE and MAE decreased, whereas the median *r*^2^ and AUC increased. This analysis revealed that the LVEF model performs well even with a single valid-class clip, but performs even better as the number of clips increases.

**Figure 4:**
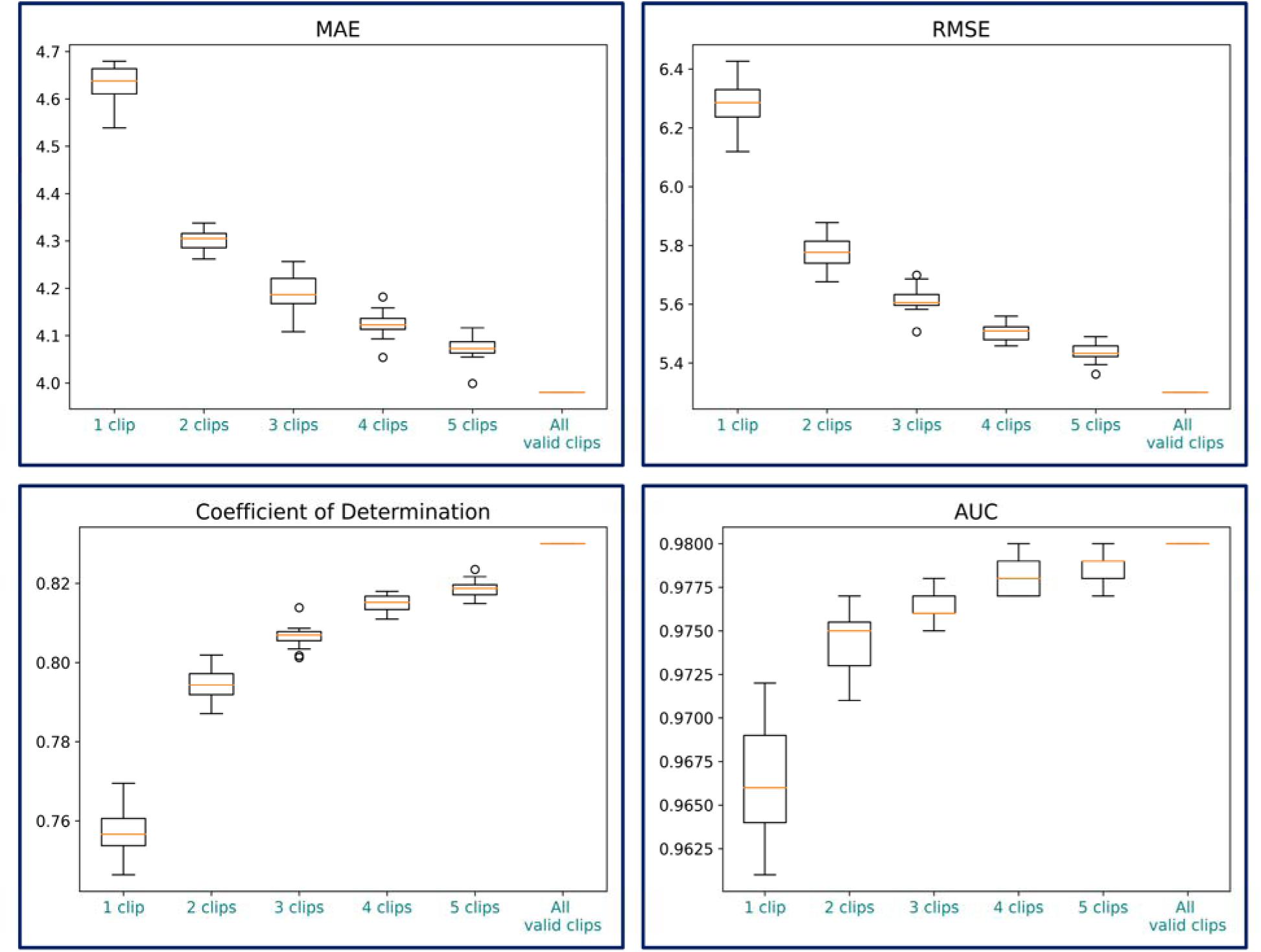
The effect of the number of clips on LVEF estimation performance for the Minnesota-Wisconsin testing dataset. The leftmost five boxplots within each panel are for one through five random clips selected from the full set of valid-class clips for a patient’s exam, whereas “All valid clips” represents all valid-class clips from a patient’s exam. In each boxplot, the center line corresponds to the median, the box limits correspond to the upper and lower quartiles, the whiskers correspond to 1.5 times the interquartile range, and the points correspond to outliers.

In addition to evaluating performance based on the number of clips (regardless of which view was selected), we also assessed model performance based on selecting specific views or combinations of views (Figure 5). Similar to the finding reported in Figure 4, this analysis revealed that model performance was generally superior when more clips were selected, but additionally revealed that when the number of views was held constant, the specific views that were taken did not have a systematic impact on performance. This finding suggests the model is indeed view-agnostic.

**Figure 5:**
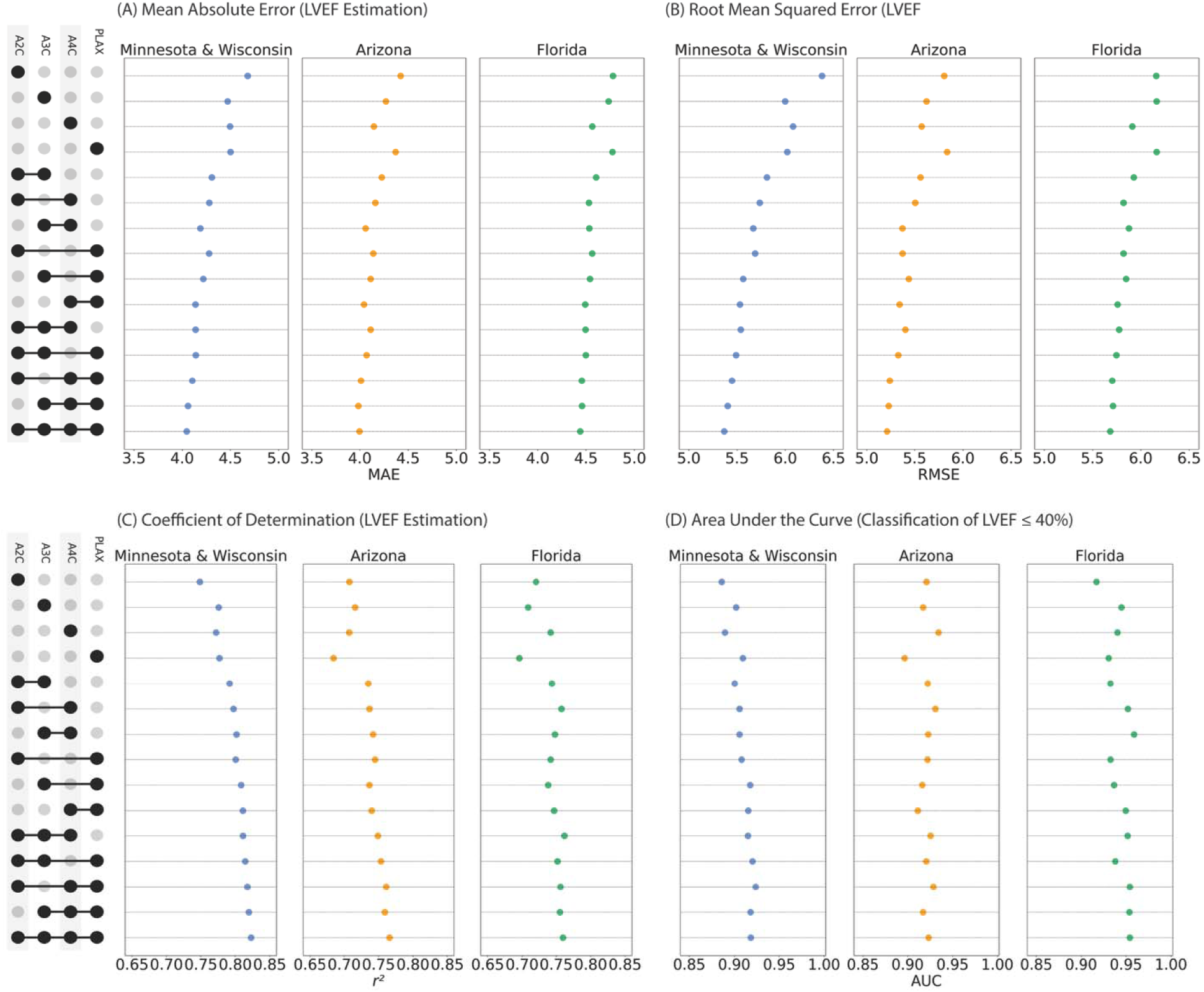
LVEF model performance for the four ‘valid’ views and combinations of these views. For combinations of views, model performance was evaluated using the average model estimate per patient across views, with only one clip taken per view (i.e., the clip with the highest score from the view classifier for that view). The legend on the left was plotted using version 0.9.0 of the *upsetplot* package ^38^ in Python version 3.9.20.

### LVEF estimation for HCU

We evaluated the LVEF regression model on HCU datasets from Mayo-Rochester and an external dataset collected by UltraSight Ltd. (Figure 6). For the Mayo-Rochester dataset, we observed very comparable results between TTE and HCU for both LVEF estimation and classification of reduced versus normal LVEF. A limitation of this experiment was that both TTE and HCU clips were collected by experienced sonographers. This limitation is addressed by the strong correlation that was observed between LVEF estimates for clips collected from expert sonographers versus novice users in the Ultrasight Ltd. dataset. In only 6% of cases, the difference in model-estimated LVEF between novice versus expert-acquired clips was greater than 10%.

**Figure 6:**
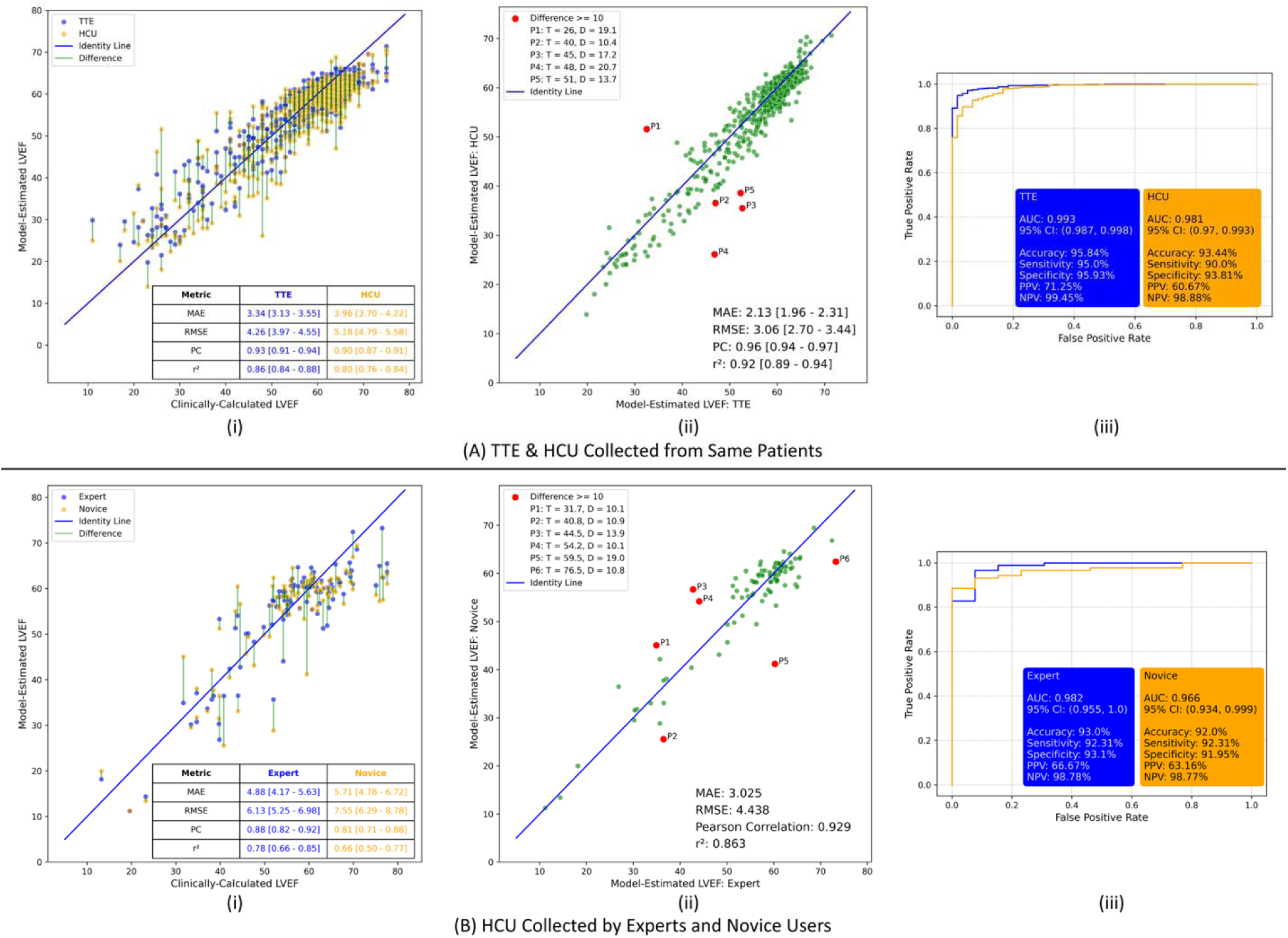
LVEF estimation and classification performance for (A) the simultaneously collected TTE-HCU dataset, and (B) the HCU dataset collected by both experts and novice users using UltraSight real-time AI guidance technology. The green lines between each set of points in the leftmost plots (i) correspond to the difference between estimates. The red points in the middle plots (ii) show the outliers for which the difference between estimates is greater than or equal to 10. For the outliers, in the legend, the true LVEF value is shown with “T” and the difference is shown with “D”. The rightmost plots (iii) show the ROC curves and metrics for reduced versus normal LVEF classification.

### Age estimation and sex classification

We observed a very strong correlation between chronological age and model-estimated age for all three of the Minnesota-Wisconsin, Arizona, and Florida cohorts, as well as a tight overlap with small differences (short green lines between each estimate pair) between AI-age estimates for HCU and TTE (Figure 7).

**Figure 7:**
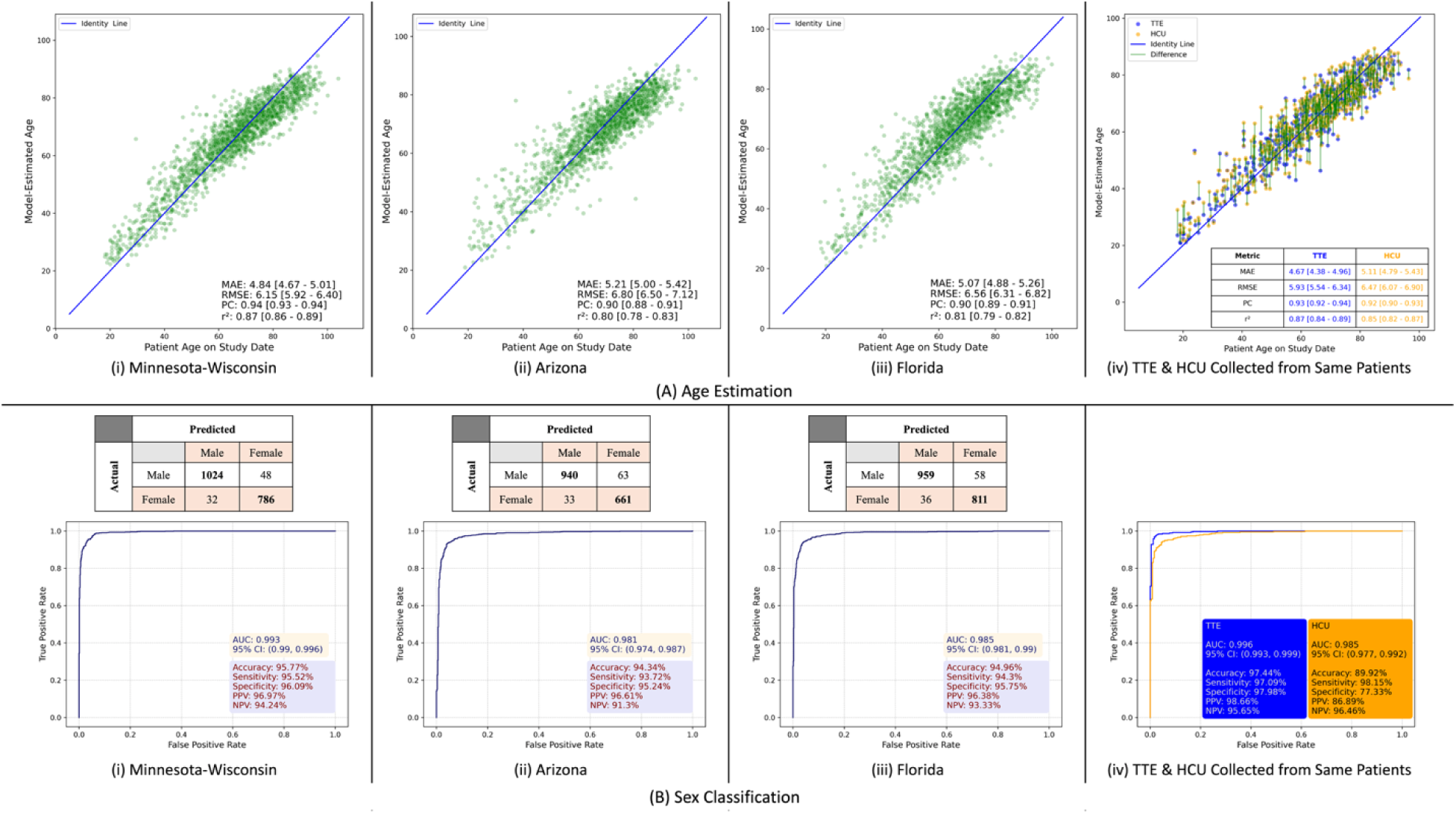
Model performance for the view-agnostic age estimation and sex classification models. The left three plots show age estimation (A) and (B) sex classification results for the TTE cohorts for both the internal (i) and external ((ii) and (iii)) testing datasets. The rightmost plot (iv) shows the overlapping correlation for HCU and TTE data collected simultaneously from the same set of patients at Mayo-Rochester, with green line between each corresponding set of estimates showing the difference between HCU versus TTE model-estimated age. The 95% confidence interval for each corresponding metric is shown in square brackets.

Sex classification model performance (Figure 8) was excellent, with an AUC greater than 0.981 for all three of the Minnesota-Wisconsin, Arizona, and Florida cohorts. Nearly overlapping ROC curves were also observed for HCU and TTE exams.

## Discussion

In this study, we created an AI framework using convolutional neural networks that are view agnostic, meaning that they do not depend on specific views to generate clinically useful results. We demonstrate that this framework can automatically derive a clinically important parameter – left ventricular ejection fraction – reliably and with a level of accuracy similar to that of human cardiologists ^10,11^. In addition, we provide evidence that this framework can also extract physiologic information beyond that which expert human readers can report (age and sex), showing the feasibility of this framework to potentially extend the capabilities of cardiac ultrasound. As we observed, this framework is not only applicable for images collected using comprehensive TTE, but also for images acquired utilizing HCU by both experienced sonographers and novice users. Consequently, the view-agnostic approach may lessen the need for user expertise in image acquisition and stands to make echocardiography a more widely available and usable clinical tool.

In the past, deep learning approaches have typically used specific views to develop and/or validate deep learning or machine learning models for LVEF estimation or classification ^2,12–26^, with some studies also segmenting the left ventricle or estimating intermediate values prior to computation of LVEF ^2,5,12–25,27–29^. However, more recent models, such as PanEcho ^30^ and DROID ^31^, estimate measurements such as LVEF without requiring segmentation or computing intermediate values, thereby eliminating potential vulnerability to errors that may arise during this process, and furthermore do not have specific view requirements, thereby allowing for greater flexibility of model input. Like PanEcho and DROID, the current set of models are also view-agnostic and do not require segmentation, but go beyond these models in their demonstrated success with HCU, as well as in their successful estimation of patient age and classification of patient sex in a view-agnostic manner.

These results have far-reaching implications. For example, rapid screening of at-risk cohorts and symptomatic patients in the clinic, emergency department, or hospital for the presence or absence of LV dysfunction would provide rapid and accurate cardiovascular triage capabilities. Additionally, these tools placed in the hands of Emergency Medical Services providers, who are typically non-expert sonographers, could allow for rapid cardiac assessment of patients before they reach the emergency department in order to better triage severe or critically ill patients prior to arrival to the hospital. Finally, although age estimation and sex classification may not have immediate clinical relevance, these tasks could potentially serve as a useful quality control measure given that ground truth values for these parameters are readily available. Furthermore, examining discrepancies between model estimates and ground truth values may allow for a better understanding of the role these factors play in cardiovascular risk prediction ^32–36^.

With that said, our work is best understood in the context of its limitations. Firstly, while the entire patient cohort was highly technically and geographically diverse, the patient cohort used for model training was not diverse in terms of race and ethnicity. This could affect the model’s accuracy and applicability across various global populations, highlighting the need for future studies to incorporate a broader patient demographic to ensure the models’ performance is universally reliable. Second, the handheld cardiac ultrasound data were collected in controlled environments, and therefore may not accurately reflect the variability typical of POCUS scenarios in diverse clinical environments. Finally, the use of a view classifier to identify valid-class views introduces the possibility of downstream errors in LVEF, age, or sex estimation.

Future work should examine model performance across a broader set of view categories than just those containing the LV, and should also consider clips collected from viewing angles outside the canonical set of views.

## Abbreviations

TTE: transthoracic echocardiography
HCU: handheld cardiac ultrasound
POCUS: point-of-care ultrasound
LVEF: left ventricular ejection fraction
ROC: receiver-operator curve
RMSE: Root Mean Square Error
MAE: Mean Absolute Error
PC: Pearson Correlation
AUC: area under the curve
CI: confidence interval(s)
PPV: positive predictive value
NPV: negative predictive value

## Data availability

All requests for raw and analysed data and related materials, excluding programming code, will be reviewed by the Mayo Clinic legal department and Mayo Clinic Ventures to verify whether the request is subject to any intellectual property or confidentiality obligations. Requests for patient-related data not included in the paper will not be considered. Any data and materials that can be shared will be released via a Material Transfer Agreement.

## Code availability

Programming code related to the PyTorch model specification will be made available under the GNU General Public License version 3 upon request to Z.I.A.

