## Supplementary Material for "A View-Agnostic Deep Learning Framework for Comprehensive Analysis of 2D-Echocardiography"

***Table of Contents***

Supplementary Methods 2

Data acquisition and selection 2

Supplementary Figure 1: Patient cohorts used for model development and evaluation 2

Model training and evaluation 5

Model selection 7

Statistics 8

References 9

### Supplementary Methods

#### Data acquisition and selection

A summary of patient cohorts is provided in Supplementary Figure 1.

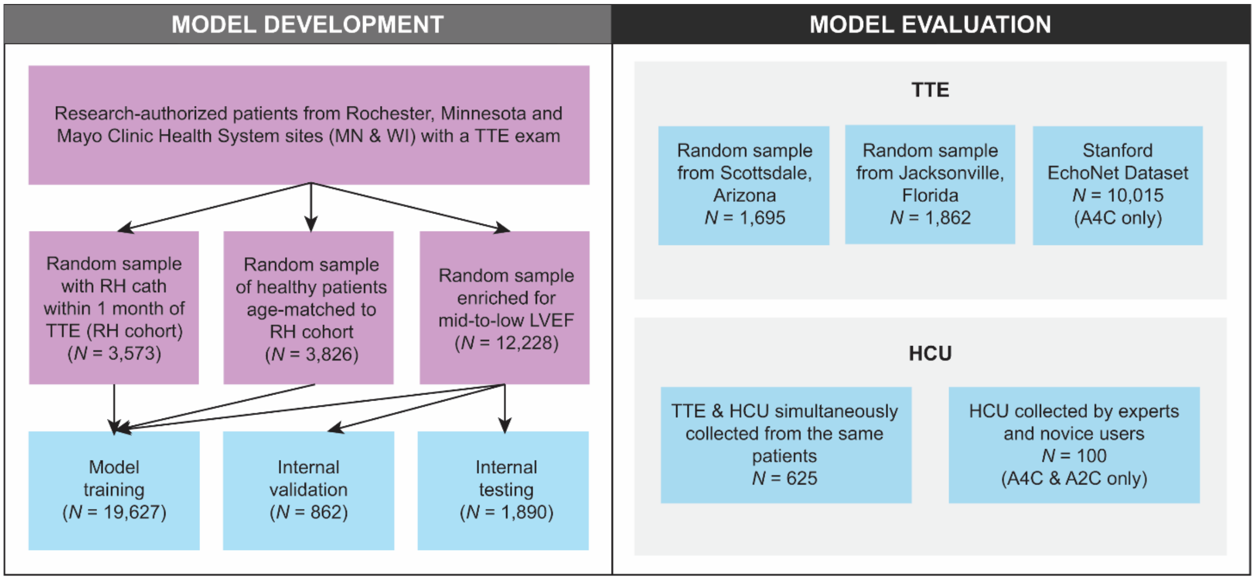


**Supplementary Figure 1:** Patient cohorts used for model development and evaluation. TTE = transthoracic echocardiography; HCU = handheld cardiac ultrasound; RH = right heart; LVEF = left ventricular ejection fraction; A4C = apical 4-chamber; A2C = apical 2-chamber

The model development cohort (one exam per patient from 19,627 patients total, with all echocardiogram study dates between January 2007 and September 2022) was an amalgamation of three distinct cohorts as follows:

1. A randomly selected sample of patients enriched for the middle to lower end of the LVEF distribution. Enrichment was performed by taking an initial entirely random sample and then supplementing it with batches of patients (randomly drawn from the list of possible studies) whose LVEF was in the following ranges: less than or equal to 40, between 41 and 45, between 46 and 50, between 51 and 55, or between 56 and 60. Exams were

selected only if they did not have more than one study phase (e.g., baseline, Valsalva), and all exams took place between January 2018 and December 2021.

2. A random selection of patients who had a diagnostic right heart catheterization procedure within a month of a TTE exam ('RH cohort'). Exams took place between January 2007 and September 2022.
3. A random selection of healthy patients who met the strict criteria of no findings of valvular disease or left or right-sided dysfunction or size abnormalities or mild or greater valvular regurgitation on any valve. Patients were age-matched to the distribution of the RH cohort. Patients with right heart strain measurements were given priority in selection (this was deemed to not impose any additional selection bias). Exams took place between March 2012 and December 2021.

For all cohorts, we excluded patients whose data were part of the training set for the view classifier embedded in our data processing workflow [1]. Then, from cohort #1 listed above, we selected groups of patients for validation and testing (not overlapping with the set of patients whose data was used for model training). Some patients were in the cohort previously used for view-specific models [2]; to ensure no data leakage, patient assignment to the training, validation, and testing cohorts was kept consistent with the previous cohort.

Inclusion and exclusion criteria for the model evaluation cohorts were as follows:

1. Arizona cohort: Exams were selected only if they did not have more than one study phase (e.g., baseline, Valsalva) and if they had an LVEF measurement belonging to the hierarchy of measurements that was used. During data processing, 145 studies (out of an

initial 1,840 studies) were excluded because they did not have at least one valid-class clip with at least 48 frames.

2. Florida cohort: Exams were selected only if they did not have more than one study phase (e.g., baseline, Valsalva) and if they had an LVEF measurement belonging to the hierarchy of measurements that was used. During data processing, 24 studies (out of an initial 1,886 studies) were excluded because they did not have at least one valid-class clip with at least 48 frames.
3. EchoNet Dynamic dataset: 15 exams were removed because clips had less than 48 frames.
4. The prospective cohort of simultaneously collected TTE and HCU data: Patients were included only if they did not belong to the model development datasets and if they had an LVEF measurement belonging to the hierarchy of measurements that was used. During data processing, 148 studies (out of an initial 773 studies) were excluded because they did not have at least one clip with at least 48 frames for both TTE and HCU for both the A2C and PLAX views. The A2C and PLAX views were used here rather than the full set of valid-class views because these are the views that were used for our previous view-specific model [2], and we wished to keep this testing set exactly the same to compare results to this previous model.
5. The set of HCU clips collected by expert sonographers and novice users: the study included 240 patients, of which 100 were selected randomly.

**Model training and evaluation**

For the training dataset, up to ten clips were randomly selected from all clips in the valid class for each patient (depending on how many were available), whereas for the testing datasets, we extracted all clips in the valid class for each patient. We randomly selected two sliding windows from each training video (passing the same sliding window twice if it had 48 frames, with different random augmentations). The rationale for the sliding window technique was that although all the videos in the model training dataset started at the peak of an R-wave, this was not the case for HCU videos in the model evaluation datasets, which instead started at a random point within the cardiac cycle. This method allowed models to be more robust to temporal variability in the cardiac cycle.

To increase model generalizability, we applied five augmentations to the training dataset: random rotation between -10 and +10 degrees, Gaussian blurring, central cropping, random cropping, and horizontal flipping [3]. We applied random rotation because we observed that due to variation in probe placement during data acquisition, the echocardiographic imaging sector (and cardiac anatomy) was sometimes not centred at the middle of each video frame; we applied Gaussian blurring to make the model more robust for lower quality echocardiographic videos; we applied central and random cropping because we observed that on some videos, some portions of the heart chambers were cropped. Gaussian blurring was performed using a 7 x 7 Gaussian kernel, and a sigma with a range from 0.1 to 0.2. Central cropping isolated the center of each frame of a video using a 180 x 180 window, with resultant images then resized back to 256 x 256 pixels. Random cropping instead isolated one of the four corners of each frame using a 180 x 180 window size, with subsequent resizing of the cropped image back to 256 x 256 pixels.

Horizontal flipping was performed so that the model would be exposed to apical 4-chamber clips in left-on-left and right-on-left orientations, since much of the training data had the left-on-left orientation used in most Mayo Clinic sites, which differs from most other sites across the world. Augmentations were performed in a batchwise fashion, with each clip having a one-sixth likelihood of either receiving no augmentation (i.e., the original data) or one of the five augmentations listed above (chosen randomly). All 24 frames within each input clip were subjected to the same augmentation. No augmentation was applied to the internal validation dataset.

When computing loss for the internal validation dataset, in each epoch, estimates per patient were obtained by first taking five randomly pulled clips with five sliding windows from each clip, passing each sliding window from each clip (i.e., 24 frames taken by sampling every other frame from a fixed-length segment) as input to the model, and then averaging model estimates across all selected windows and clips for that patient. Trained models were saved, and the best models were selected based on minimum patient-wise loss for the validation dataset.

All models were developed using the Python programming language and PyTorch framework [4]. The LVEF estimation model was trained for 80 epochs for 7 days, the age estimation model was trained for 58 epochs for 6 days, and the sex classification model was trained for 100 epochs for 6 days using four Tesla A100 GPUs, each with 40GB of memory. The root mean square error (RMSE) loss function was used during model training for the LVEF and age estimation models, whereas a cross-entropy loss function was used for sex classification. A batch size of 40 was used, divided equally across four GPUs using the data parallelization technique. We used a learning rate of 0.001 and optimization was performed using the Adam algorithm [5] with weight decay factor of 0.00001. We also used the mixed precision technique [6] for faster and more balanced model training.

For model evaluation, the LVEF estimate for each exam was the average of the model output from up to ten (depending on the number of available frames) overlapping sliding windows (e.g., every other frame from frames 0 to 47, 12 to 59, 24 to 71, all the way up to frames 108 to 155) for all available valid-class clips for that exam. For age, the evaluation process was exactly as described for LVEF, whereas for sex classification, we averaged the probability scores for up to ten sliding windows, with the final classification decision based on the average probability score.

**Model selection**

The S3D model architecture was selected out of a set of six candidate model architectures that we used to train a set of internal models on a subset of the current training dataset (2100 patients). The candidate model architectures that we evaluated were: 3DCNN (view-agnostic version of our previously developed view-specific model [2]), ResNet Mixed Convolution (MC3) [7], ResNet 18 (3D) [7], ResNet (2+1)D [7], S3D [8], and Video Swin Transformer [9]. Taking both performance and computational costs into account, we selected S3D as our model architecture. More details about the S3D model architecture can be found in the original manuscript [8]. Note that for the LVEF and age estimation models, we passed 1 as the argument value for number of classes, whereas for sex classification, we passed 2 as the argument value.

**Statistics**

To evaluate model performance for LVEF and age estimation (i.e., continuous regression), we computed RMSE, mean absolute error (MAE), the Pearson correlation (PC) coefficient (r), and the coefficient of determination (r2). 95% confidence intervals (CI) for all four metrics (RMSE, MAE, PC, and r2) were calculated using bootstrapping [10] with 10,000 resamples and a fixed random seed of 100. For LVEF classification and sex classification, we computed accuracy, sensitivity, specificity, positive predictive value (PPV), negative predictive value (NPV), and area under the receiver operating characteristic curve (AUROC, abbreviated as AUC), with a 95% CI. We used the fast implementation of DeLong's method [11] to compute 95% CIs for AUCs. Detailed explanations regarding many of these metrics can be found at [12,13].
